# Estimation of real-infection and immunity against SARS-CoV-2 in Indian populations

**DOI:** 10.1101/2021.02.05.21251118

**Authors:** Prajjval Pratap Singh, Rakesh Tamang, Manoj Shukla, Abhishek Pathak, Anshika Srivastava, Pranav Gupta, Alay Bhatt, Abhishek K. Shrivastava, Sudhir K. Upadhyay, Ashish Singh, Sanjeev Maurya, Purnendu Saxena, Vanya Singh, Akhilesh Kumar Chaubey, Dinesh Kumar Mishra, Yashvant Patel, Rudra Kumar Pandey, Ankit Srivastava, Nargis Khanam, Debashruti Das, Audditiya Bandopadhyay, Urgyan Chorol, Nagarjuna Pasupuleti, Sachin Kumar Shrivastav, Satya Prakash, Astha Mishra, Pavan Kumar Dubey, Ajit Parihar, Priyoneel Basu, Jaison J Sequeira, KC Lavanya, Vijayalaxmi, Bhat. K. Vishnu Shreekara, Thadiyan Parambil Ijinu, Dau Dayal Aggarwal, Anand Prakash, Kiran Yadav, Anupam Yadav, Vandana Upadhyay, Gunjan Mukim, Ankan Bhandari, Ankita Ghosh, Akash Kumar, Vijay Kumar Yadav, Kriti Nigam, Abhimanyu Harshey, Tanurup Das, Deepa Devadas, Surendra Pratap Mishra, Ashish, Abhay Kumar Yadav, Nitish Kumar Singh, Manpreet Kaur, Sanjay Kumar, Nikhil Srivastava, Charu Sharma, Ritabrata Chowdhury, Dharmendra Jain, Abhai Kumar, Ritesh Shukla, Raghav Kumar Mishra, Royana Singh, Yamini B Tripathi, Vijay Nath Mishra, Mohammed S. Mustak, Niraj Rai, Sumit Kumar Rawat, Prashanth Survajhala, Keshav K Singh, Chandana Basu Mallick, Pankaj Shrivastava, Gyaneshwer Chaubey

**Author notes:** Prajjval Pratap Singh, Rakesh Tamang, Manoj Shukla and Abhishek Pathak - equal first author contribution.

## Abstract

Infection born by Coronavirus SARS-CoV-2 has swept the world within a time of a few months. It has created a devastating effect on humanity with social and economic depressions. Europe and America were the hardest hit continents. India has also lost several lives, making the country fourth most deadly worldwide. However, the infection and death rate per million and the case fatality ratio in India were substantially lower than many of the developed nations. Several factors have been proposed including the genetics. One of the important facts is that a large chunk of Indian population is asymptomatic to the SARS-CoV-2 infection. Thus, the real infection in India is much higher than the reported number of cases. Therefore, the majority of people are already immune in the country. To understand the dynamics of real infection as well as level of immunity against SARS-CoV-2, we have performed antibody testing (serosurveillance) in the urban region of fourteen Indian districts encompassing six states. In our survey, the seroprevalence frequency varied between 0.01-0.48, suggesting high variability of viral transmission among states. We also found out that the cases reported by the Government were several fold lower than the real infection. This discrepancy is majorly driven by a higher number of asymptomatic cases. Overall, we suggest that with the high level of immunity developed against SARS-CoV-2 in the majority of the districts, it is less likely to have a second wave in India.

## Introduction

The whole world has been fighting the COVID-19 epidemic for over a year. During this time, the number of people who died due to infection of this virus in the globe has also crossed a mark of 2 millions (Dong et al., 2020). Before the roll-out of vaccination recently, it was highly challenging for a nation to stop the spread of this virus. Several of the standard precautionary measures e.g. strict lockdown, use of mask, frequent hand hygiene, social distancing, contact tracing and quarantine have been applied widely (Jung et al., 2020; Rodriguez-Palacios et al., 2020). Even after this, no country is yet in such a condition that it can declare itself completely free from it. Initially, herd immunity was also considered and discussed widely (Randolph and Barreiro, 2020; Slot et al., 2020).

Herd immunity against the SARS-CoV-2 is one of the highly debated topic (Anderson and May, 1985; Bock et al., 2020; Frederiksen et al., 2020; Jung et al., 2020; Kwok et al., 2020; Neagu, 2020). During the pandemic when approximately 70% of the population infect and develop antibodies against the pathogen, it facilitates the herd immunity. This barrier of immunity blocks the virus and adds passive immunity to non-infected people. However, due to the high fatality rate of SARS-CoV-2 infection, nearly all of the countries did not adopt this strategy. For example, United Kingdom (UK) and Sweden have opted where UK abandoning it at the initial stage (Hunter, 2020; Sibony, 2020), whereas Sweden was heavily criticised with largest number of cases and fatalities in Scandinavia (Habib, 2020; Orlowski and Goldsmith, 2020). Thus, the plan of obtaining a rapid herd immunity was indeed an autophagy (Chaubey, 2020).

The model of herd immunity is very similar to the positive selection (or selective sweep), a term often used in the genomic studies (Sabeti et al., 2002; Voight et al., 2006). Similarly, we can also relate hard sweep and soft sweep terminology (Pritchard et al., 2010), for a soft and hard herd immunity. With the exclusion of hard herd immunity, the alternate option was to obtain the soft herd immunity with strict use of defensive procedures. Protective measures like strict lockdown, use of masks, frequent hand hygiene and social distancing have significantly reduced the effective infections and overburden to the emergency services (Aleta et al., 2020; West et al., 2020). Also with the efficacious contact tracing and quarantine, one may achieve soft herd immunity with period of time (Aleta et al., 2020).

From the first case in India on 30^th^ Jan 2020, now the total cases have been surpassed to >10 million with >0.15 million fatalities (Coronavirus in India: Latest Map and Case Count). Although still the new number of cases are between 10-15,000 per day, India has followed a perfect bell-shaped curve with a well-defined peak in September 2020. Several studies have pinpointed that the real number of infected people in a population are several fold higher than the reported cases (Aspelund et al., 2020; Böhning et al., 2020; Ivorra et al., 2020; Mukhopadhyay and Chakraborty, 2020; Pedersen and Meneghini, 2020; Shaman, 2020). In addition with that, a large number of asymptomatic people add another layer of complexity to it which have been seen in case of India (Chaubey, 2020). Most of the studies relied upon the mathematical modelling which may not show the real picture of COVID-19 in India ((Arti and Bhatnagar, 2020; Chatterjee et al., 2020a, 2020b; Mukhopadhyay and Chakraborty, 2020; Tiwari, 2020). The random antibody testing in a population is one of the procedures to obtain the real-time picture of the developed immunity. Thus, to understand the COVID-19 dynamics in India, we have performed the real-time antibody testing on urban populations from fourteen districts of India. The aim was to estimate the level of immunity against COVID-19 among urban street vendors.

### Materials and Methods

Since most of the COVID-19 cases in India were concentrated on urban areas, we have selected urban populations in our survey. During the month of September-December 2020, we randomly surveyed healthy working individuals who have neither been diagnosed with COVID-19, nor had been sick with any associated symptom in the recent past. Moreover, we excluded those individuals in our survey whose family members have been ever diagnosed with the COVID-19 in the past. Our focus was to screen people from urban vendor group who are relatively more exposed in the society e.g. roadside workers, roadside fruit-vegetable sellers, rikshaw pullers, autorickshaw drivers, milkman, hawkers etc. With these stringent criteria, we screened unrelated people who have not lived together.

We have used Coviscreen™ kit, kindly provided by Biosense Technologies, India to screen the individuals for the presence of antibodies. This kit detects the total antibody (IgM+IgA+IgG) present in the blood. We followed the manufacturers protocol for the detection. A sample test for positive and negative samples have been shown in Supplementary Figure 1. Complete details about this kit can be obtained from the supplier http://www.tulipgroup.com/Covid19/Covid19.html.

The frequency of Antibody positive individuals were calculated in each of the districts surveyed (Table 1). The districtwise geographical frequency map was generated by https://www.datawrapper.de/. Frequency bar plot of each district was drawn with 95%CI (Figure 1). District-wise population census data was extracted from the 2011 census of India (Census of India Websitel⍰: Office of the Registrar General & Census Commissioner, India). Numbers of reported cases in each district were recorded from the Covid India tracker (Coronavirus in India: Latest Map and Case Count). This study has been approved by the Institutional Ethical Committee of Banaras Hindu University, Varanasi, India.

**Figure 1.**
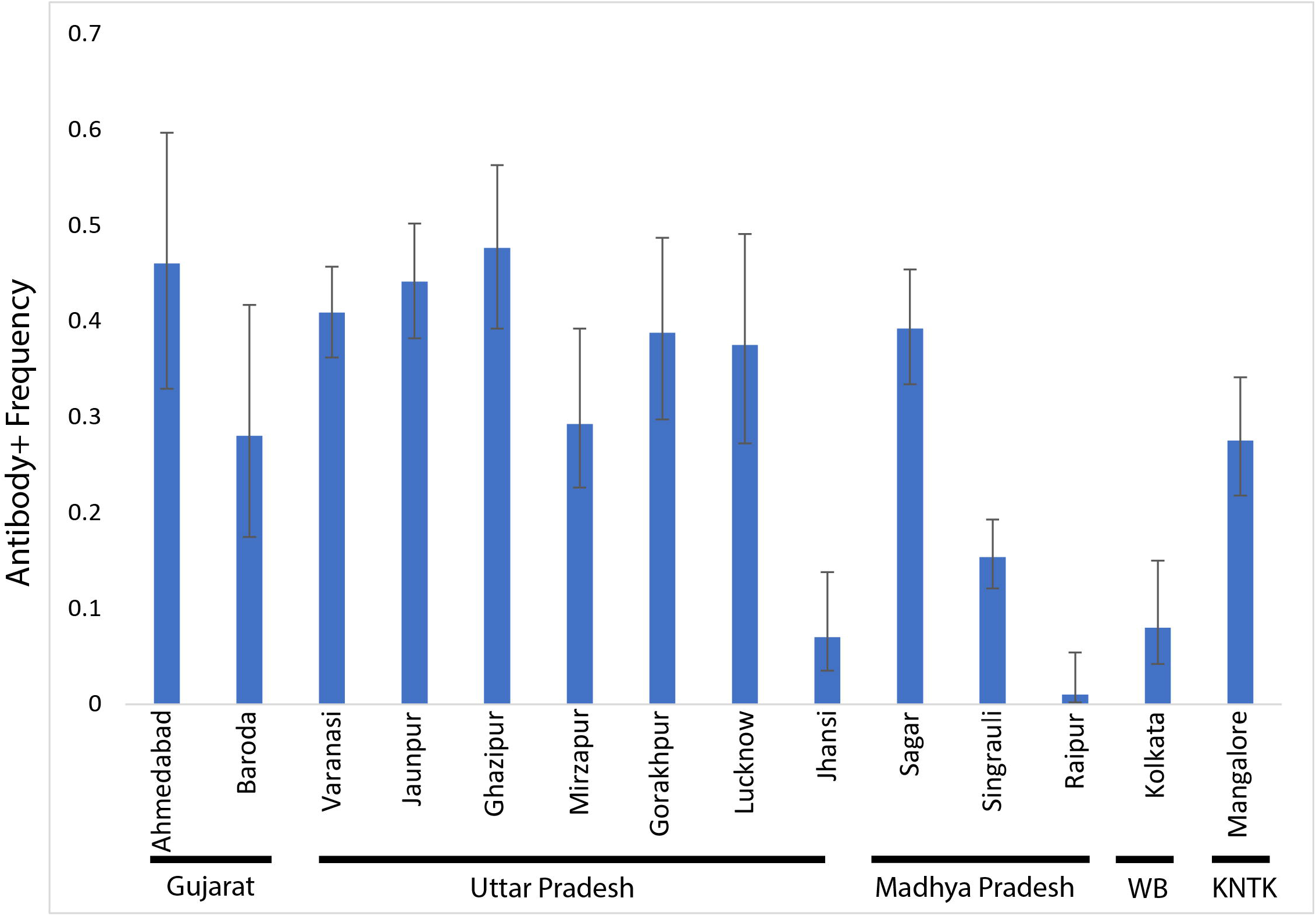
The bar plot showing seroprevalence frequency (CI95%) among studied districts. The districts are also grouped in to states.

**Table 1.** The detailed districtwise data of seroprevalence. Frequency of Government cases has been calculated by number of reported cases/urban census. Number of times were calculated by dividing seroprevalence with frequency of Government reported cases.

| State | District | Months | N | Antibody+ | Freq.(Ab+) | Freq.(Govt.) | Cases (Govt.) | Urban Census | Times |
| --- | --- | --- | --- | --- | --- | --- | --- | --- | --- |
| Gujarat | Ahmedabad | Dec-20 | 50 | 23 | 0.46 (0.329-0.597) | 0.011 | 60848 | 5600000 | 42.3 |
| Gujarat | Baroda | Dec-20 | 50 | 14 | 0.28 (0.175-0.417) | 0.017 | 27755 | 1671000 | 16.9 |
| Uttar Pradesh | Varanasi | Sept 2020-Dec 2020 | 406 | 166 | 0.409 (0.362-0.457) | 0.012 | 23360 | 2000000 | 35.0 |
| Uttar Pradesh | Jaunpur | Sept 2020-Dec 2020 | 263 | 116 | 0.441 (0.382-0.502) | 0.019 | 6697 | 347000 | 22.9 |
| Uttar Pradesh | Ghazipur | Sept 2020-Dec 2020 | 128 | 61 | 0.477 (0.392-0.563) | 0.043 | 5236 | 121000 | 11.0 |
| Uttar Pradesh | Mirzapur | Oct-20 | 100 | 30 | 0.30 (0.219-0.396) | 0.014 | 3441 | 246000 | 21.4 |
| Uttar Pradesh | Gorakhpur | Oct-20 | 98 | 38 | 0.388 (0.297-0.487) | 0.031 | 21364 | 695000 | 12.6 |
| Uttar Pradesh | Lucknow | Oct-20 | 72 | 27 | 0.375 (0.272-0.491) | 0.029 | 81276 | 2817000 | 13.0 |
| Uttar Pradesh | Jhansi | Nov-20 | 100 | 7 | 0.07 (0.035-0.138) | 0.019 | 10399 | 548000 | 3.7 |
| Madhya Pradesh | Sagar | Sept 2020-Dec 2020 | 250 | 98 | 0.392 (0.334-0.454) | 0.015 | 5367 | 370000 | 27.0 |
| Madhya Pradesh | Singrauli | Sept 2020-Oct 2020 | 384 | 59 | 0.154 (0.121-0.193) | 0.008 | 1907 | 227000 | 18.3 |
| Chhattishgarh | Raipur | Sep-20 | 100 | 1 | 0.01 (0.002-0.054) | 0.047 | 53106 | 1124000 | 0.2 |
| West Bengal | Kolkata | Dec-20 | 100 | 8 | 0.08 (0.042-0.15) | 0.009 | 127656 | 14036000 | 8.8 |
| Karnataka | Mangalore | Dec-20 | 200 | 55 | 0.275 (0.218-0.341) | 0.054 | 33701 | 624000 | 5.1 |

## Results and Discussion

In our survey, geographically we have covered districts from Western, Northern, Eastern, Central and Southern regions of India. Two districts were covered from Western part (Gujarat), seven districts were surveyed in Northern part (Uttar Pradesh), three districts from Central part (Madhya Pradesh and Chhattisgarh), whereas single district each of Eastern (West Bengal) and Southern (Karnataka) regions were surveyed (Table 1). Our seroprevalence survey in fourteen districts of six states have shown high level of variability among the regions (Supplementary Fig. 2). The minimum antibody positive people were observed in Raipur district of Chhattisgarh state, whereas maximum antibody positive individuals were found in Ghazipur district of Uttar Pradesh state (Figure 1 and Table 1). The mean seroprevalence among studied districts was 0.306 (95%CI 0.287-0.325). The high prevalence of antibodies in many of the districts suggests that the soft herd immunity in India is underway. The frequency of antibody positive people is sporadic, however regions like Eastern Uttar Pradesh showed largely a uniform seroprevalence (Fig. 1 and Supplementary Fig. 2). The mean value in this region was found to be 0.41 (95%CI 0.38-0.44). Major districts of this region like Varanasi, Jaunpur and Ghazipur all had a frequency of >0.40 seroprevalence. The variation among districts suggests that the level of virus spread among districts of India is asymmetrical.

Notably, all the participants in this study were found to be asymptomatic, they did not know that they ever had COVID-19. This suggests that a large proportion of Indian population is asymptomatic against SARS-CoV-2 infection. Indeed, there might be several known and unknown rationales behind that, however the prevalence of rs2285666 associated haplotype (Srivastava et al., 2020) and 35 years or younger age of ∼65% of Indian populations, are two highly likely explanations.

In order to understand the magnitude of unreported cases, we have compared the frequency of government reported cases with the frequency of antibody positive cases. All of the districts showed higher seroprevalence than reported cases (Table 1). The mean value of studied districts was 17 times (95% CI 13-22) higher than the government reported cases. Similar with the frequency dissimilarities, in statewise comparison, we observed substantial differences e.g. 40 times in Gujarat, 21 times in Uttar Pradesh, 23 times in Madhya Pradesh, 0.2 times in Chhattisgarh, 9 times in West Bengal and 5 times in Karnataka. Interestingly, we found out that cases reported by the Government were highly correlated with the urban census (two tailed p<0.0001), whereas we did not find any significant correlation between antibody positive frequency with urban census or government reported cases.

As our result suggests that the number of real infections in India is several fold higher than the Government reported cases, we may reconsider the case fatality ratio. The present case fatality ratio of India (1.44) is significantly lower (two tailed p value <0.0001) than global average (2.15). Considering our result which shows a far greater number of real infected people, the actual case fatality ratio of India is at least 17 times lower (Table 1).

Our observation at the fourteen districts of India indicates sporadic high level of immunity among people who are highly contagious as well as exposed. Since our focus was on the people who are more prone to receive as well as spread infection. Our result advocates that a strong level of immunity wall has been already in effect. Keeping in mind such a scenario it is likely that most of the hotspots are saturated with the immune people. This is why the number of daily infections are decreasing and the chance of a second wave is highly unlikely.

In summary, for the first time we have used a novel approach to estimate the developed immunity against SARS-CoV-2 among the exposed population of fourteen districts. These results strongly indicate that the soft herd immunity is already in force in most of the Indian territory. We observed that on an average every third street vendor in majority of the studied districts is immune against SARS-CoV-2. The number of asymptomatic cases in India is much higher than expected. Overall, we suggest that with the high level of immunity developed against SARS-CoV-2 in the majority of districts, it is unlikely to have a second wave in India.

## Data Availability

All the data is available with the manuscript.

## Acknowledgements

We are grateful to the Biosense Technologies, India for their kind help in this project. We did not receive any specific funding for this project.

## Data Availability Statement

All datasets generated for this study are included in the article/Supplementary Material.

## Figure Legend

**Supplementary Figure 1**. The detection of antibodies by the Coviscreen™ kit. The kit produce one and two bands in case of Antibody negative and positive persons respectively.

**Supplementary Figure 2**. The geospatial frequency of antibodies with our sampling locations.

