## Supplementary figures and images for "Estimation of real-infection and immunity against SARS-CoV-2 in Indian populations"

### Supplementary Fig. 1

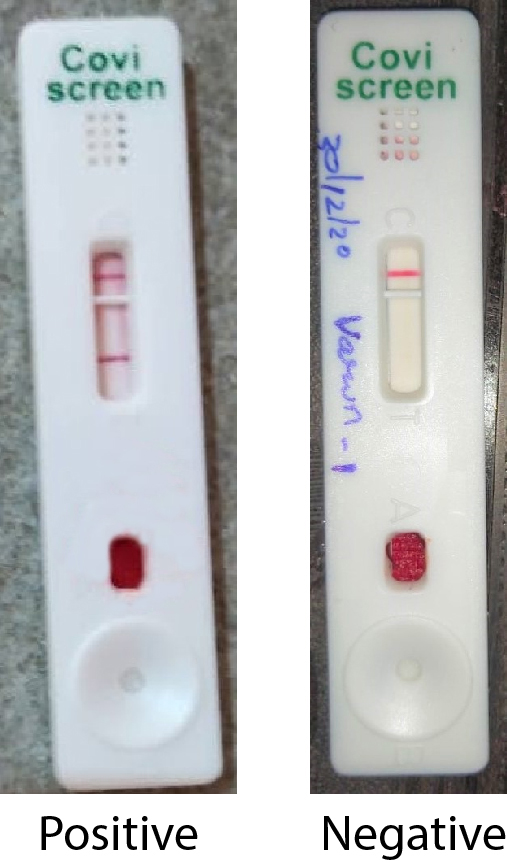

### Supplementary Figure 2

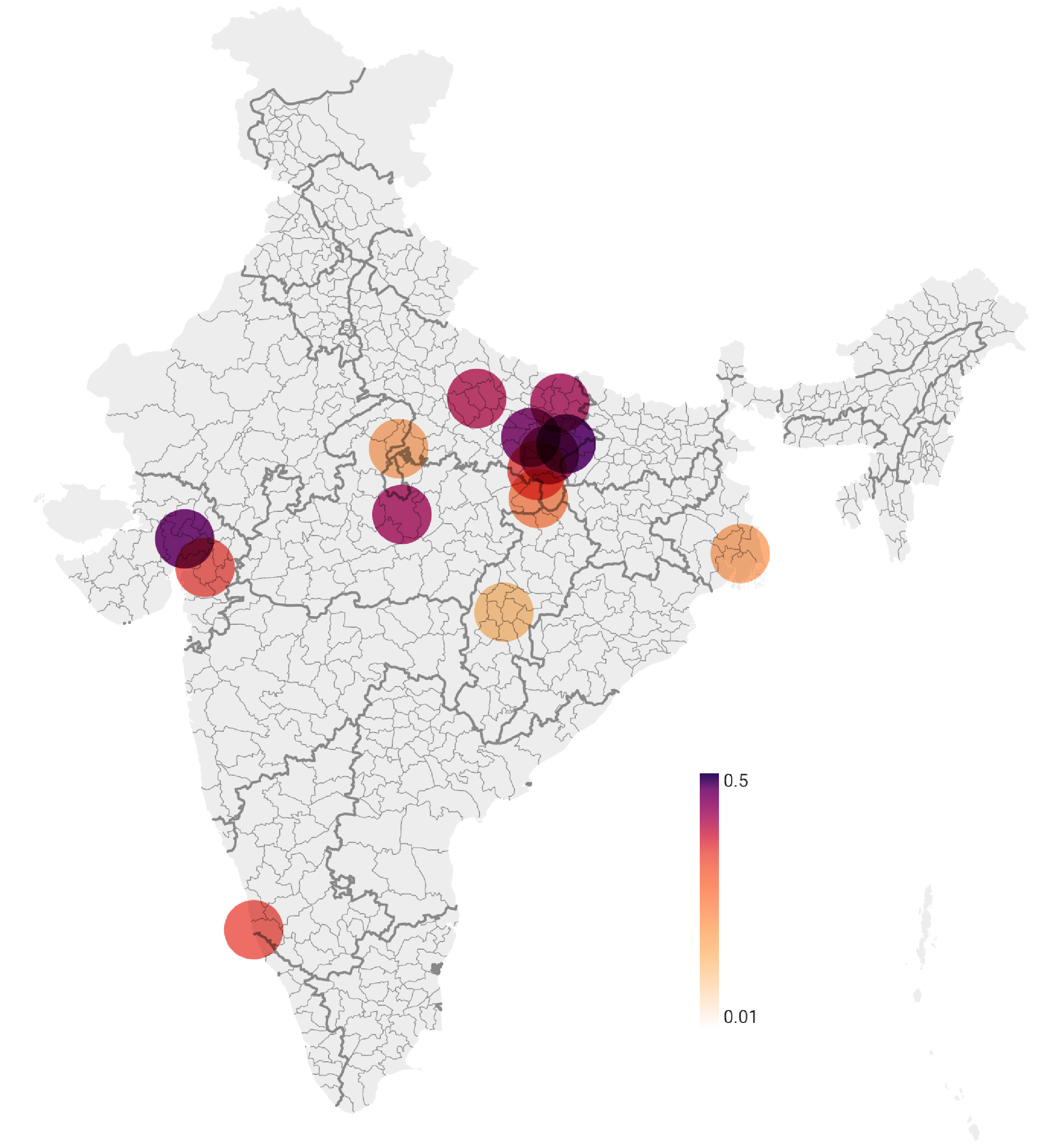
